# Neutralizing antibodies correlate with protection from SARS-CoV-2 in humans during a fishery vessel outbreak with high attack rate

**DOI:** 10.1101/2020.08.13.20173161

**Authors:** Amin Addetia, Katharine HD Crawford, Adam Dingens, Haiying Zhu, Pavitra Roychoudhury, Meei-Li Huang, Keith R. Jerome, Jesse D. Bloom, Alexander L. Greninger

**Affiliations:** Department of Laboratory Medicine and Pathology, University of Washington School of Medicine, Seattle, WA; Division of Basic Sciences and Computational Biology Program, Fred Hutchinson Cancer Research Center, Seattle, WA; Department of Genome Sciences, University of Washington, Seattle, WA; Medical Scientist Training Program, University of Washington, Seattle, WA; Vaccine and Infectious Disease Division, Fred Hutchinson Cancer Research Center, Seattle, WA; Howard Hughes Medical Institute, Seattle, WA

## Abstract

The development of vaccines against SARS-CoV-2 would be greatly facilitated by the identification of immunological correlates of protection in humans. However, to date, studies on protective immunity have only been performed in animal models and correlates of protection have not been established in humans. Here, we describe an outbreak of SARS-CoV-2 on a fishing vessel associated with a high attack rate. Predeparture serological and viral RT-PCR testing along with repeat testing after return to shore was available for 120 of the 122 persons on board over a median follow-up of 32.5 days (range 18.8 to 50.5 days). A total of 104 individuals had an RT-PCR positive viral test with Ct <35 or seroconverted during the follow-up period, yielding an attack rate on board of 85.2% (104/122 individuals). Metagenomic sequencing of 39 viral genomes suggested the outbreak originated largely from a single viral clade. Only three crewmembers tested seropositive prior to the boat’s departure in initial serological screening and also had neutralizing and spike-reactive antibodies in follow-up assays. None of these crewmembers with neutralizing antibody titers showed evidence of bona fide viral infection or experienced any symptoms during the viral outbreak. Therefore, the presence of neutralizing antibodies from prior infection was significantly associated with protection against re-infection (Fisher’s exact test, p=0.002).

## Introduction

Severe acute respiratory syndrome coronavirus 2 (SARS-CoV-2) has caused tens of millions of infections and hundreds of thousands of deaths worldwide since its emergence in December 2019. Multiple vaccine candidates are currently in Phase III trials (1-3). The success of these vaccines could be helped by further insights into the protective nature of neutralizing antibodies in humans.

Neutralizing antibodies have been isolated from individuals previously infected with SARS-CoV-2 (4, 5). These antibodies often target the receptor binding domain (RBD) of the SARS-CoV-2 spike (S) protein and prevent the binding interaction between the spike protein and the host’s angiotensin-converting enzyme 2 (ACE2) (4, 5), although neutralizing antibodies that do not inhibit spike’s binding to ACE2 have also been identified (6, 7). In animal models, neutralizing antibodies are protective against SARS-CoV-2 (8, 9).

Vaccines currently in development against SARS-CoV-2 have been shown to elicit levels of neutralizing antibodies comparable to those observed in naturally infected persons (1-3). However, the protective nature of both vaccine- and infection-elicited neutralizing antibodies in humans remains unproven, with animal models being used to make inferences about protection (10, 11). Human challenge trials, which could provide rapid information about the protection conferred by neutralizing antibodies (12, 13), are controversial due to the severity and unknown long-term impacts of SARS-CoV-2 infection and concerns over ethical administration of such trials (14, 15).

Given the high number of people exposed to SARS-CoV-2 every day, retrospective analyses of outbreak events may provide insights into the protective nature of neutralizing antibodies. In particular, outbreaks on confined shipping vessels are particularly useful candidates for assessing protection from SARS-CoV-2 infection (16-18). The high population density and large degree of contact between people on ships contributes to a high attack rate. In some cases nearly all passengers will have been exposed (16).

Here, we performed a retrospective analysis of a SARS-CoV-2 outbreak on a fishing vessel that departed from Seattle, Washington in May 2020. Predeparture viral and serological testing was performed on the near entirety of the ship’s crew, allowing for testing of how pre-existing immunity correlated with subsequent infection during the outbreak.

## Methods

### Clinical diagnostic testing

Nasopharyngeal swabs were collected from patients in 3 mL of viral transport media. RT-PCR testing was performed on either the Hologic Panther Fusion, Roche cobas 6800, or the University of Washington CDC-based, emergency use authorized laboratory developed test (19). Clinical testing of serum samples was performed using the Abbott Architect SARS-CoV-2 IgG assay (20). Index values associated with the Abbott test are chemiluminescent signal values relative to a calibrator control, and are broadly similar to O.D. values for an ELISA. An index value ≥ 1.40 is qualitatively reported as positive. The case definition for an individual infected on the boat included anyone with a positive RT-PCR with Ct < 35 or seroconversion by the Abbott test during the follow-up period. This study was approved by the University of Washington Institutional Review Board.

### SARS-CoV-2 whole genome sequencing

RNA was extracted from positive SARS-CoV-2 samples using the Roche MagNA Pure 96 (21). Metagenomic sequencing libraries were constructed as previously described (22). Briefly, RNA was DNAse-treated using the Turbo DNA-Free Kit (Thermo Fisher). First strand cDNA was synthesized using Superscript IV (Thermo Fisher) and 2.5μM random hexamers (IDT) and second strand synthesis was performed with Sequenase Version 2.0 DNA Polymerase (Thermo Fisher). The resulting double-stranded cDNA was purified using 1.6X volumes of AMPure XP beads (Beckman Coulter). Libraries were constructed using the Nextera DNA Flex Pre-Enrichment kit (Illumina) and cleaned using 0.7X volumes of AMPure XP beads. The resulting libraries were sequenced on a 1×75 bp Illumina NextSeq run. A median of 509,551 sequencing reads were obtained for each sample. Sequencing reads are available at NCBI BioProject PRJNA610428 and sequence accessions are available in Supplemental Table 1.

Consensus genomes were called using a custom SARS-CoV-2 genome calling pipeline (https://github.com/proychou/hCoV19). Briefly, sequencing reads were adapter- and quality-trimmed with BBDuk and mapped to the SARS-CoV-2 reference genome (NC_045512.2) using Bowtie 2 (23). Reads aligning to the SARS-CoV-2 reference genome were filtered using BBDuk and assembled with SPAdes (24). The *de novo* assembled contigs and mapped read assemblies were merged to produce a consensus genome. For samples that did not produce a genome through the automated pipeline, the mapped read assemblies were visualized in Geneious and a consensus genome was called manually.

A phylogenetic analysis was completed using the 39 consensus genomes obtained through metagenomic sequencing and 109 other SARS-CoV-2 isolates downloaded from https://www.gisaid.org/ (accessed July 17, 2020) reflective of the global genomic diversity of SARS-CoV-2. To select 109 SARS-CoV-2 isolates, all global SARS-CoV-2 sequences were downloaded from GISAID. Those composed of >5% Ns, those with disrupted reading frames, and those with partial genomes were discarded. The strains were then stratified by Pangolin lineage (A or B) (https://github.com/cov-lineages/pangolin) and 49 from lineage A and 59 from lineage B were randomly selected along with the Wuhan-Hu-1 reference genome (NC_045512.2) (25). Sequences were aligned with MAFFT v7.453 (26) and a phylogenetic tree was constructed using FastTree (version 2.1.1) (27) with the 5’ and 3’UTRs masked. The resulting phylogenetic tree was visualized in R (version 3.6.1) using the ggtree package (28). Strains most closely related to the major outbreak clade were identified by searching against a custom BLASTN database containing all SARS-CoV-2 sequences in GISAID (accessed August 3, 2020).

### Neutralization Assays and Anti-Spike Antibody Testing

The presence of anti-Spike and neutralizing antibodies was analyzed in pre-departure sera samples from individuals that were positive in the Abbott assay screening through four different methods: Spike IgG ELISA, RBD ELISA, ACE2 blockade of binding ELISA, and pseudovirus neutralization.

RBD and spike protein for the ELISAs were produced as described previously (29). IgG enzyme-linked immunosorbent assays (ELISAs) to spike and RBD were adapted from published protocol (30, 31), with details described previously (32). Spike or RBD was diluted to 2 μg/mL in PBS and 50 μL/well was used to coat 96 well Immunlon 2HB plates (Thermo Fisher; 3455) at 4°C overnight. Plates were washed three times the next day with PBS containing 0.1% Tween 20 (PBS-T) using a Tecan HydroFlex plate washer. Plates were blocked for 1 hour with 200 μL/well of 3% non-fat dry milk in PBS-T at room temperature. Sera were diluted 4-fold in PBS-T containing 1% non-fat dry milk, starting at a 1:25 dilution. Pooled sera collected from 2017-2018 from 75 individuals (Gemini Biosciences, 100-110, lot H86W03J) and CR3022 antibody (starting at 1/ug/mL, also diluted 4-fold) were included as negative and positive controls, respectively. After block was thrown off plates, 100μL diluted sera was added to plates and incubated at room temperature for 2 hours. Plates were again washed three times, and then 50μL of a 1:300 dilution of goat anti-human IgG-Fc horseradish peroxidase (HRP)-conjugated antibody (Bethyl Labs, A80-104P) in PBS-T containing 1% milk was added to each well and incubated for 1 hour at room temperature. Plates were again washed three times with PBS-T. 100μL of TMB/E HRP substrate (Millipore Sigma; ES001) was then added to each well, and after a 5-minute incubation, 100 μL 1N HCl was added to stop the reaction. 0D450 values were read immediately on a Tecan infinite M1000Pro plate reader. Area under the titration curve (AUC) was calculated with the dilutions on a log-scale.

The ACE2 blockage of binding assay was performed using the SARS-CoV-2 Surrogate Virus Neutralization Test Kit (GenScript). The assay was performed following the manufacturer’s recommendations with 10μL serum diluted into 90μL dilution buffer and read using the DS2 microplate reader (Dynex technologies).

Neutralization assays with spike-pseudotyped lentiviral particles were performed as described previously (33), with a few modifications. Briefly, cells were seeded in black-walled, clear bottom, poly-L-lysine coated 96-well plates (Greiner, 655936). About 14 hours later, serum samples were diluted in D10 media (DMEM with 10% heat-inactivated FBS, 2 mM l-glutamine, 100 U/mL penicillin, and 100 μg/mL streptomycin) starting with a 1:20 dilution followed by 6 serial 3-fold dilutions. An equal volume of full-length spike-pseudotyped lentiviral particles as diluted serum was added to the serum dilutions and incubated at 37C for 1 hour. 100μL of the virus plus serum dilutions were then added to the cells ~16 hours after the cells were seeded.

About 52 hours post-infection, luciferase activity was measured as described previously (33) except luciferase activity was read out directly in the assay plates without transferring to black, opaque bottom plates. Two “no serum” wells were included in each row of the neutralization plate and fraction infectivity was calculated by dividing the luciferase readings from the wells with serum by the average of the “no serum” wells in the same row. After calculating the fraction infectivity, we used the neutcurve Python package (https://jbloomlab.github.io/neutcurve/) to calculate the serum dilution that inhibited infection by 50% (IC50) by fitting a Hill curve with the bottom fixed at 0 and the top fixed at 1. All serum samples were measured in duplicate. To calibrate our neutralization assays, we also ran them on the NIBSC reference serum sample (product number 20/130) and measured an IC50 of 1:2395.

## Results

### Predeparture PCR and serology testing

There were a total of 122 people (113 men and 9 women) on the manifest of the ship. Prior to the ship’s departure, crewmembers were screened for active SARS-CoV-2 infection by RT-PCR, or for serological evidence of prior or ongoing infection using the Abbott Architect assay which detects antibodies against the viral nucleoprotein (N). Predeparture RT-PCR and serology test data were available for 120 crewmembers. This predeparture screening occurred on Day 0 and Day 1 prior to the ship’s departure on Day 2. In this predeparture screening, none of the crewmembers tested positive for virus by RT-PCR, and six individuals tested seropositive in the Abbott Architect assay (index value ≥1.40) (Figure 1 A).

**Figure 1.**
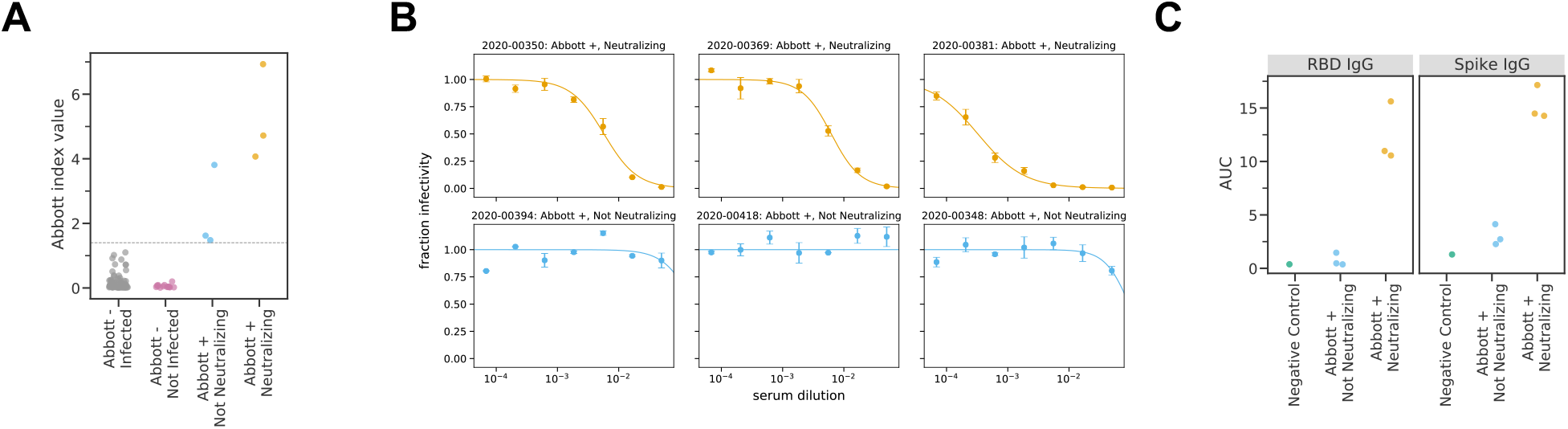
Pre-departure serological assays. A) Abbott Architect index values for all 120 individuals assayed. The grey line indicates the cutoff for a positive Abbott reading (≥ 1.40). Individuals with negative Abbott index values are further classified by whether they subsequently became infected on the ship. Individuals with positive Abbott index values are further characterized by whether their pre-boarding serum was neutralizing. B) Neutralization curves for all 6 pre-boarding samples that were positive in the Abbott Architect assay. C) Titers of RBD- or Spike-binding IgG antibodies in all 6 Abbott positive pre-boarding samples as measured by ELISA. The negative control sample is pooled sera collected in 2017-2018 from 75 individuals (Gemini Biosciences, 100-110, lot H86W03J).

**Table 1.** Laboratory values for crew members who were pre-departure seropositive by Abbott SARS-CoV-2

| Sample | Day 0-1 |  |  |  |  |  | Day 18-21<br>PCR (Ct) | Day 25-26<br>PCR | Day 28<br>PCR | Day 31-36<br>PCR | Day 31-35<br>Abbott IgG index | Day 31-35<br>ACE2 BoB |
| --- | --- | --- | --- | --- | --- | --- | --- | --- | --- | --- | --- | --- |
|  | RT-PCR | Abbott IgG<br>index | Neutralization<br>IC50 | ACE2<br>BoB | RBD IgG<br>AUC | spike IgG<br>AUC |  |  |  |  |  |  |
| 2020-00350 | negative | 6.93 | 1:174 | 89% | 15.62 | 17.15 | negative | negative | n.d. | negative | 6.40 | 95% |
| 2020-00369 | negative | 4.07 | 1:161 | 84% | 10.98 | 14.27 | negative | n.d. | n.d. | negative | 2.93 | 68% |
| 2020-00381 | negative | 4.72 | 1:3082 | 93% | 10.56 | 14.48 | negative | 37.4 | negative | 38.3 | 3.48 | 90% |
| 2020-00394 | negative | 1.62 | >1:20 | -4% | 1.46 | 4.13 | 22.91 | n.d. | n.d. | 27.9 | 4.29 | 30% |
| 2020-00418 | negative | 3.81 | >1:20 | 3% | 0.47 | 2.27 | 22.84 | n.d. | n.d. | 30.4 | 6.31 | 93% |
| 2020-00348 | negative | 1.48 | >1:20 | 0% | 0.37 | 2.72 | 17.57 | n.d. | n.d. | negative | 5.98 | 35% |

n.d., not done; BoB, blockade of binding

After becoming aware of the subsequent SARS-CoV-2 outbreak on the ship (see next section), we tested residual predeparture serum samples from the six individuals who were seropositive in the Abbott Architect assay to characterize the neutralizing and spike-binding activity of their sera. The sera of three of these six individuals had potent neutralizing activity against SARS-CoV-2 spike pseudotyped lentiviral particles (Table 1, Figure 1B). The neutralizing titers (1:174, 1:161, 1:3082) are in the typical range of titers observed in humans who have been infected with SARS-CoV-2 within the previous few months (29, 34, 35). The sera of the three individuals with neutralizing titers also had high activity in an assay that measure the ability of antibodies to block RBD binding to ACE2, as well as in IgG ELISAs against spike and RBD (Table 1, Figure 1C).

Notably, the sera of the other three individuals who were seropositive in the Abbott Architect assay but did not have neutralizing activity had lower quantitative readings in the Abbott assay (including two that were close to the cutoff of 1.40; Figure 1A) and readings comparable to those from negative controls in the RBD and spike ELISA assays (Figure 1C). Therefore, we speculate that the three individuals without neutralizing activity were false positives in the initial serological screening. However, they could have been in the early stages of active infection, since the Abbott Architect detects antibodies against N while all the other assays we used detect antibodies against spike, and anti-N antibodies appear earlier after infection than anti-spike antibodies (36, 37). Alternatively, they could have experienced a mild or asymptomatic infection, which can be associated with transient or low-level seroconversion (38, 39).

Overall, assuming that only individuals who were positive in the initial Abbott Architect assay have neutralizing anti-spike antibodies, then just three of the 120 individuals with pre-departure screening data had neutralizing antibodies prior to boarding the ship. We consider this assumption to be well supported by several lines of evidence: large-scale studies have demonstrated that the Abbott Architect has close to 100% sensitivity by two weeks post-symptom onset (20); numerous studies (36, 37) have shown that SARS-CoV-2 infected patients almost invariably mount strong and early antibody responses to the N antigen detected by the Abbott Architect; and a study (32) using the exact assays described here found that only individuals with anti-N antibodies have neutralizing titers to SARS-CoV-2.

### Testing after ship returned due to outbreak

On Day 18, the ship returned to shore after a crewmember became sick, tested positive for SARS-CoV-2, and required hospitalization. Testing data after return was available for all 122 crewmembers for RT-PCR and 114 crewmembers for serology using the Abbott assay. RT-PCR and serological testing was performed until day 50, leading to a median follow-up of 32.5 days (range 18.8 to 50.5 days).

Of the 118 individuals with RT-PCR results from the week of return, 98 tested positive with a Ct < 35. Three additional crewmembers tested positive by RT-PCR with a Ct < 35 within the next 10 days. The median of the strongest/minimum Ct for each of these 101 crewmembers who tested positive with Ct < 35 was 22.8 (IQR 19.3 - 26.9). Serological responses among these individuals as measured by Abbott SARS-CoV-2 IgG index value increased for the majority of these individuals (Figure 2A).

**Figure 2.**
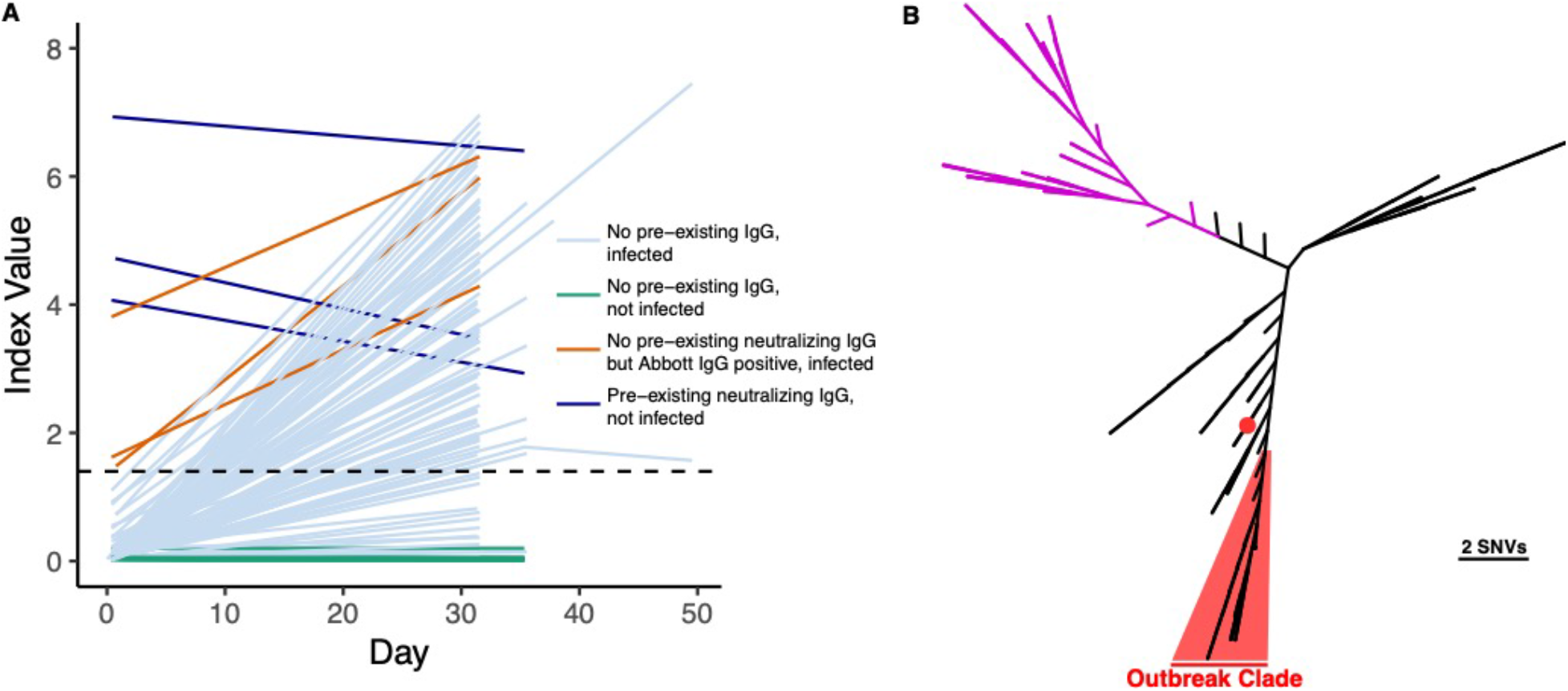
Return to shore testing. A) Abbott Architect SARS-CoV-2 index values over time (pre- and post-departure) are depicted for each individual with at least 2 serum draws. The dashed line denotes the seropositivity cutoff of the assay (1.40). Individuals who had a positive RT-PCR with Ct < 35 or who seroconverted during the follow-up period are shown in light blue. Individuals who were not infected by the above case definition criteria are shown in green. Individuals who screened positive by the Abbott Architect SARS-CoV-2 IgG assay but lacked neutralizing antibodies and were infected are shown in brown. Individuals who had pre-existing neutralizing antibodies and were not infected are shown in blue. B) SARS-CoV-2 whole genome sequencing of cases from the fishery vessel confirms outbreak. SARS-CoV-2 genomes from 39 cases with Ct < 26 were recovered and a phylogenetic tree was made using FastTree along with 109 other isolates reflective of global diversity. 38 cases are highlighted in red with a median pairwise difference of 1 single nucleotide variant, while one outlier case from the boat is shown with a red dot. Clade A strains associated with early trans-Pacific transmission are shown in purple.

Among the 21 crewmembers who never had a positive RT-PCR test with Ct < 35, three individuals seroconverted based on Abbott Architect index value during the follow-up period. Two of these three crewmembers had positive RT-PCR values with Ct values > 35, while RT-PCR data was not available for the third until Day 49. These three individuals were considered infected on the vessel. In addition, three of the 21 crewmembers without a positive RT-PCR result with Ct < 35 were not tested by serology after returning to shore, though two of the three crewmembers tested negative 3 and 4 times, respectively, by RT-PCR over three weeks after returning.

### Confirmation of outbreak with whole genome sequencing

Metagenomic recovery of 39 SARS-CoV-2 whole genomes from the outbreak indicated a major single outbreak clade (FastTree support value: 1.00) covering 38 isolates that differed by a median of one nucleotide across the genome (range 0-5) (Figure 2B). Sixteen of these isolates shared completely identical sequence. The closest SARS-CoV-2 whole genome sequences in GISAID (August 3, 2020) to the major outbreak clade were strains from Virginia (USA/VA-DCLS-0561/2020), New York City (USA/NY-NYUMC650, NYUMC624, NYNYUMC474, NYUMC426/2020), Minnesota (USA/MN-MDH-1288/2020), or Michigan (USA/MI-MDHHS-SC20223/2020) at 2 SNVs apart.

### The three crewmembers with neutralizing antibodies were protected from infection

We can assess the effects of pre-existing neutralizing antibodies on infection during the outbreak using the pre-departure serological screening (available for 120 of 122 individuals) and the subsequent testing of all 122 individuals for infection. None of the three individuals who had neutralizing antibodies prior to departure were infected during the subsequent outbreak using our case definition of a positive RT-PCR test with Ct < 35 or seroconversion, and none reported any symptoms upon return to shore. In contrast, among the other 117 of 120 individuals with pre-departure serological data who were seronegative or lacked spike-reactive antibodies prior to departure, 103 of 117 were infected using the same case definition (of the 2 individuals without pre-departure serological screening, one tested positive and one tested negative by RT-PCR on return). Therefore, the overall rate of infection was 0 of 3 among individuals with neutralizing antibodies, and 103 of 117 among individuals without such antibodies.

This difference is statistically significant (Table 2, Fisher’s exact test P = 0.002), indicating that pre-existing neutralizing antibodies are significantly associated with protection against SARS-CoV-2 infection.

**Table 2.** Summary table of infection status of crew members for which pre-departure serology testing was performed.

|  |  | Pre-departure |  |
| --- | --- | --- | --- |
|  |  | Neutralizing<br>Ab (+) | Neutralizing<br>Ab (-) |
| On boat | Infected | 0 | 103 |
|  | Not Infected | 3 | 14 |

$p=0.0024$

The three crewmembers who were seropositive for anti-N antibodies by Abbott but did not have neutralizing antibodies were all infected during follow-up, with minimum Cts of 17.6, 22.8, and 22.9 and increases in Abbott index values (Table 1). Sex did not differ between uninfected and infected, with females composing 5.6% (1 of 18) and 7.7% (8 of 104) of these two groups, respectively (Fisher’s exact test, p=1).

We also looked in detail at the viral testing results of the three crewmembers who were positive for neutralizing antibodies to assess the strength of the evidence that they were not re-infected during this ship outbreak. Two tested fully negative by RT-PCR on 3+ occasions, with negative tests on Days 18, 25, 35, and 36 and Days 18, 35, and 36. The third individual tested negative on the Roche cobas on Day 21 and Day 28, and positive only by the E-gene primers/probe set (Ct 37.4) and negative by the orf1ab primer set on the Roche cobas on Day 25. This individual also tested positive (Ct 38.3) on Day 31 on the Hologic Panther Fusion. By our case definition (which required a positive RT-PCR test with Ct < 35), these results are not consistent with being infected on the boat. The sporadic high-Ct results could be consistent with intermittent, low-level shedding associated with recent past infection, as low levels of SARS-CoV-2 have been detected in nasal passages for more than 80 days (40). Of note, only two other crewmembers had a minimum Ct > 35 in the post-departure follow-up period and both of these individuals were considered infected due to seroconversion during the follow-up period. In contrast, Abbott index values decreased for all three of the crewmembers with predeparture neutralizing antibodies during the follow-up period.

## Discussion

Here, we report an outbreak of SARS-CoV-2 on a fishing vessel with an attack rate greater than 85%. Screening with the Abbott Architect anti-nucleocapsid IgG antibody test followed by confirmation of positives with multiple anti-spike protein antibody tests including neutralization assays demonstrated the protective nature of neutralizing antibodies. In particular, none of the three individuals with pre-existing neutralizing antibodies were infected, whereas the vast majority of other individuals were infected. These findings are consistent with data from animal models, in which the elicitation of high titers of neutralizing antibodies was protective against re-challenge with SARS-CoV-2 (8, 10, 41).

An assumption of our analysis is that the only individuals who had pre-existing neutralizing and anti-spike antibodies were those who tested seropositive in the initial pre-departure Abbot Architect anti-N serological screening, since only individuals positive in that screening were subjected to additional serological assays for anti-spike and neutralizing antibodies. However, this assumption is well supported by the validated high sensitivity of the Abbott Architect assay (20), plus the well-established fact that anti-N antibodies appear earlier that anti-spike antibodies (36, 37). Additionally, our four anti-spike antibody tests showed a high level of consistency among seropositive samples, and prior work using the exact same assays has found neutralizing antibodies only among individuals who were positive in the Abbott Architect assay (32). As shown by others, the RBD ELISA and neutralizing antibody assays were highly consistent (42, 43). The ACE2 blockade of binding functional ELISA assay showed excellent consistency with the more laborious pseudovirus neutralizing antibody assay (44).

It is intriguing that one individual who had predeparture neutralizing antibodies and was classified as uninfected by our case definition nonetheless had a sporadic very weak signal in viral testing on two different RT-PCR platforms. It is well-established that SARS-CoV-2 can be detected for multiple weeks in the nasopharyngeal tract, well after the resolution of symptoms and elicitation of an antiviral immune response (45, 46). However, it is unclear at this time whether immunity to SARS-CoV-2 will be sterilizing (10, 47), and it is possible that the sporadic weak signal in viral testing for this individual was the result of re-exposure to virus on the boat.

In prior studies, the Abbott SARS-CoV-2 IgG assay has shown excellent performance characteristics with high specificity (99.1-99.9%) for prior infection with SARS-CoV-2 (20, 48, 49). Curiously, the positive predictive value for the Abbott SARS-CoV-2 IgG assay for neutralizing antibodies or protection in our population was only 50% (3/6 crewmembers). It is difficult to conclusively determine whether these represented false positives or just anti-N/anti-spike discrepants, particularly given that anti-N antibodies tend to appear before anti-spike antibodies (36, 37). All three of the individuals who were Abbott IgG positive prior to departure but lacked neutralizing and anti-spike antibodies and were RT-PCR positive upon return showed strong increases in index value. In addition, two of these three individuals had pre-departure Abbott index values that were close to the positivity cut-off. Unfortunately, we did not have sufficient residual pre-departure serum to run on a separate anti-N platform such as the Roche Elecsys anti-SARS-CoV-2 (50).

This study is limited by lack of information on clinical symptoms for the majority of crewmembers on the vessel and direct knowledge of contacts on the boat. We cannot also necessarily know that the three individuals with neutralizing antibodies prior to departure were exposed directly to SARS-CoV-2 on the vessel. The study is also limited by the low seroprevalence in the predeparture cohort—which is consistent with the approximate seroprevalence in May 2020 in the Seattle area, but means that there were only three individuals with pre-existing neutralizing antibodies. Nonetheless, with an overall attack rate of >85%, the lack of infection in the three individuals with neutralizing antibodies was statistically significant compared to the rest of the boat’s crew. Overall, our results provide the first direct evidence anti-SARS-CoV-2 neutralizing antibodies are protective against SARS-CoV-2 infection in humans.

## Data Availability

Data in supplementary tables

## Acknowledgements

The authors thank Nicole Lieberman for helpful comments and Nathan Breit for data pulls. We also thank Brooke Fiala, Samuel Wrenn, Deleah Pettie, and Neil P. King at the Institute for Protein Design for sharing reagents for ELISA assays. Work performed in the clinical laboratory was supported by the Department of Laboratory Medicine and Pathology. This research was supported by the following grants from the NIAID of the NIH: R01AI141707 (to J.D.B.) and F30AI149928 (to K.H.D.C.). J.D.B. is an Investigator of the Howard Hughes Medical Institute.

---

**Supplemental Table 1.** SARS-617 CoV-2 isolates and accessions sequenced in this study.

| Isolate | GISAIID Accession Number |
| --- | --- |
| USA/WA-UW-10027/2020 | EPI_ISL_461450 |
| USA/WA-UW-10028/2020 | EPI_ISL_461451 |
| USA/WA-UW-10029/2020 | EPI_ISL_461452 |
| USA/WA-UW-10030/2020 | EPI_ISL_511852 |
| USA/WA-UW-10031/2020 | EPI_ISL_461453 |
| USA/WA-UW-10034/2020 | EPI_ISL_511853 |
| USA/WA-UW-10036/2020 | EPI_ISL_461454 |
| USA/WA-UW-10038/2020 | EPI_ISL_511854 |
| USA/WA-UW-10039/2020 | EPI_ISL_461455 |
| USA/WA-UW-10040/2020 | EPI_ISL_461456 |
| USA/WA-UW-10042/2020 | EPI_ISL_461457 |
| USA/WA-UW-10088/2020 | EPI_ISL_461458 |
| USA/WA-UW-10089/2020 | EPI_ISL_461459 |
| USA/WA-UW-10090/2020 | EPI_ISL_461460 |
| USA/WA-UW-10091/2020 | EPI_ISL_461461 |
| USA/WA-UW-10093/2020 | EPI_ISL_461462 |
| USA/WA-UW-10094/2020 | EPI_ISL_461463 |
| USA/WA-UW-10101/2020 | EPI_ISL_511855 |
| USA/WA-UW-10102/2020 | EPI_ISL_461464 |
| USA/WA-UW-10105/2020 | EPI_ISL_511856 |
| USA/WA-UW-10106/2020 | EPI_ISL_461465 |
| USA/WA-UW-10107/2020 | EPI_ISL_461466 |
| USA/WA-UW-10108/2020 | EPI_ISL_461467 |
| USA/WA-UW-10113/2020 | EPI_ISL_511857 |
| USA/WA-UW-10114/2020 | EPI_ISL_461468 |
| USA/WA-UW-10115/2020 | EPI_ISL_511858 |
| USA/WA-UW-10116/2020 | EPI_ISL_512086 |
| USA/WA-UW-10117/2020 | EPI_ISL_461469 |
| USA/WA-UW-10118/2020 | EPI_ISL_461470 |
| USA/WA-UW-10124/2020 | EPI_ISL_511859 |
| USA/WA-UW-10126/2020 | EPI_ISL_511860 |
| USA/WA-UW-10127/2020 | EPI_ISL_461471 |
| USA/WA-UW-10128/2020 | EPI_ISL_461472 |
| USA/WA-UW-10129/2020 | EPI_ISL_461473 |
| USA/WA-UW-10130/2020 | EPI_ISL_461474 |
| USA/WA-UW-10131/2020 | EPI_ISL_461475 |
| USA/WA-UW-10133/2020 | EPI_ISL_511861 |
| USA/WA-UW-10136/2020 | EPI_ISL_461476 |
| USA/WA-UW-10138/2020 | EPI_ISL_461477 |

